# Cardiovascular-Kidney-Metabolic Syndrome, Social Determinants of Health, and Dementia Risk: a prospective cohort study

**DOI:** 10.1101/2025.03.17.25324154

**Authors:** Hui Zhang, Zixin Hu, Xiangnan Li, Yi Li, Xiaofeng Wang, Li Jin, Shuai Jiang, Xiangwei Li, Meng Hao

## Abstract

Cardiovascular-kidney-metabolic (CKM) syndrome, characterized by pathophysiological interactions among metabolic risk factors, chronic kidney disease and the cardiovascular system, is a significant global health concern, particularly in populations with adverse social determinants of health (SDOH). However, the influence of CKM syndrome and the joint effects of SDOH on the risk of developing incident dementia has not been fully elucidated. Here, we examined these associations among 382569 individuals from the UK Biobank. We found that unfavorable SDOH and advanced CKM syndrome (stage 3-4) are independently associated with increased risks of incident all-cause dementia, Alzheimer’s disease and vascular dementia. Additionally, joint associations of CKM syndrome stages and SODH with incident dementia were observed. Individuals with ≥3 unfavorable SDOH and in CKM syndrome stage 4 exhibited the highest dementia risk, even after adjusting for APOE ε4 status. Our findings highlighted the importance of maintaining optimal CKM health and addressing unfavorable SODH in cognitive aging.

## Introduction

Dementia poses a significant and growing global public health concern, affecting approximately 57 million people in 2019 and projected to rise to 153 million by 2050 due to population ageing and growth^1^. It is estimated that up to 47.0%-72.6% of dementia cases could be preventable through modifications to lifestyle, local environments, and social factors^2–5^. Notably, emerging evidence of epidemiological studies indicates a significantly higher risk of incident dementia among patients with many chronic diseases, including diabetes, cardiovascular disease (CVD), chronic kidney disease (CKD), and metabolic syndrome^2–4^. Additionally, CVD, CKD, and metabolic syndrome are deeply intertwined, and frequently coexisted^6–8^. They often share pathophysiological mechanisms and risk factors^8–10^. In response, the American Heart Association (AHA) has recently introduced the concept of cardiovascular-kidney-metabolic (CKM) syndrome, characterized by pathophysiological interactions among metabolic risk factors, CKD and the cardiovascular system^11^. According to the AHA, CKM syndrome was classified into five stages (0-4) to indicate its risk spectrum, and manifested as a holistic and systemic health disorder that impacted nearly all organ systems and contributed to multiorgan diseases^11^. Therefore, considering the intricate interplay within CKM function, it is particularly meaningful to identify and intervene on dementia-related risk factors from a holistic perspective of CKM health.

At present, observational studies have demonstrated significant association of CKM syndrome with increased risks of chronic diseases and mortality across diverse populations^12–15^. More importantly, several national surveys reported a widespread prevalence of CKM syndrome in China^16^, U.S^17^, and South Korea^18^. For example, approximately 15% of adults met criteria for advanced CKM syndrome (stages 3-4) in general US population^17^. Notably, an especially higher prevalence of advanced CKM syndrome was also observed in individuals with unfavorable social determinants of health (SDOH), such as unemployed, food insecurity, and lower educational level^17–20^. In summary, elucidating the relationship between CKM syndrome stages and dementia risk is imperative for optimizing risk stratification, facilitating early preventive interventions, and improving prognosis. However, the relationships between CKM syndrome, SDOH, and their combined impact on incident dementia risk has not been fully elucidated. To date, few studies have investigated these potential relationships in prospective cohort studies.

To address the identified knowledge gaps, we first posited that CKM syndrome would be associated with an increased risk of incident dementia, with SDOH significantly influencing this relationship. Subsequently, we performed a longitudinal analysis using UK Biobank data to explore the potential associations of individual CKM syndrome, SDOH, and their interactions with dementia risk.

## Methods

### Study population

The UK Biobank is a large-scale, population-based, and prospective cohort study. It consisted of over 500000 adults aged 37-73 years recruited across England, Scotland, and Wales. During the baseline period (2006 to 2010), questionnaires, physical measurements, and biological samples were used to obtain information about individuals. All individuals provided electronic informed consent. The UKB study received approval from the National Information Governance Board for Health and Social Care and the National Health Service North West Multi-Centre Research Ethics Committee. In this study, a total of 382569 individuals with complete data on both CKM syndrome, SDOH, and dementia were included and analyzed.

### Assessment of CKM syndrome

In UK Biobank, body mass index (BMI), waist circumstance, systolic and diastolic blood pressure (SBP, DBP) were measured. Fasting blood samples were collected for laboratory tests, including total cholesterol, high-density and low-density lipoprotein cholesterol (HDL, LDL), triglyceride, fasting blood glucose, glycosylated hemoglobin, cystatin C, and creatinine were measured. In this study, metabolic syndrome is defined by the presence of ≥3 of the following: (1) waist circumference: ≥88 cm for women and ≥102 cm for men (if Asian ancestry, ≥80 cm for women and ≥90 cm for men), (2) HDL <40 mg/dL for men and <50 mg/dL for women; (3) triglycerides ≥150 mg/dL; (4) blood pressure: SBP ≥130 mmHg and/or DBP ≥80 mmHg and/or use of antihypertensive medications; and (5) fasting blood glucose ≥100 mg/dL. CKD was defined as estimated glomerular filtration rate (eGFR) of <60 mL/min/1.73 m^2^, or a urinary albumin-creatinine ratio (UACR) of >30 mg/g at enrollment, or a history of CKD^21^. The eGFR based on both serum creatinine and cystatin C was calculated using the race-free CKD-EPI 2021 creatinine equation^21^. Individuals with eGFR ranged from 45 to 60 ml/min/1.73m^2^ were recognized as moderate-to-high-risk CKD, eGFR ranged from 15 ml/min/1.73m^2^ to <45 ml/min/1.73m^2^ were recognized as very-high-risk CKD, respectively.

According to the AHA’s definition^22^, five stages of CKM syndrome were defined: stage 0 (no CKM risk factors), stage 1 (excess and/or dysfunctional adiposity), stage 2 (metabolic risk factors or CKD), stage 3 (subclinical CVD in CKM) and stage 4 (clinical CVD in CKM). In detail, Stage 0 was characterized by the absence of CKM risk factors. The stage included individuals without overweight/obesity (BMI ≥25 kg/m^2^; if Asian ancestry ≥23 kg/m^2^), abdominal obesity (waist circumference ≥88/102 cm in women/men; if Asian ancestry ≥80/90 cm), hypertension (self-reported history of hypertension, current antihypertensive treatment, SBP ≥140 mmHg, and/or DBP ≥90 mmHg), diabetes (self-reported history of diabetes, current hypoglycemic treatment, or fasting blood glucose 124 mg/dL, or glycated hemoglobin ≥6.5%), metabolic syndrome, and hypertriglyceridemia (triglyceride ≥135 mg/dL), CKD, or subclinical/clinical CVD. Stage 1 included individuals with overweight/obesity, abdominal obesity, dysfunctional adiposity (fasting blood glucose level between 100 mg/dL and 124 mg/dL, or glycated hemoglobin level between 5.7% and <6.5%); but without the presence of other metabolic risk factors, or CKD, or subclinical/clinical CVD. Stage 2 included individuals with metabolic risk factors, or CKD, but without subclinical/clinical CVD. Stage 3 included individuals with both subclinical CVD and CKM risk factors (excess/dysfunctional adiposity, other metabolic risk factors, or CKD). Subclinical CVD was defined as having high predicted 10-year CVD risk. High 10-year CVD risk was defined as a 10-year risk of at least 10% as estimated using the AHA Predicting Risk of CVD EVENTs (PREVENT) equations by the AHA^23–25^. Stage 4 was identified based on self-reported established CVD (coronary heart disease, angina, heart attack, heart failure, and stroke) and CKM risk factors.

### Social determinants of health

The SDOH of interest included nine items, and were dichotomized into favorable vs unfavorable conditions according to the conventional cut points. Unfavorable SDOH was assessed by self-reported questionnaires, included low educational levels, lower income, unemployment, not owning a home, living alone, fewer social support, less social/leisure activity, less social connection, emotional distress^26,27^. The detail assessments were presented in **Supplemental Table 1**. Low income was defined as if their total household income was less than £31000. Participants were asked their employed situation, those not working were classified as unemployment, except students or retired persons. Housing ownership was determined by asking if the home they were living in was owned or being bought, rented, or occupied by some other arrangement. Not owning a home was defined if the accommodation that you live in is rent. Living status was assessed by the number of people who living together in household. Living alone was defines if they answered 1 people. Social support was assessed by the frequency of confide in someone close, individuals who answered never or almost never were defined as social isolation. Social activity/leisure was assessed by the frequency of attending social/leisure activity. Individuals who did not attend any group activities were defined as less social/leisure activity. Social connection was assessed by the frequency of visiting each other among friends/family, Individuals who answered about or less once a month were defined as less social connection. Emotional distress was defined if individuals have experienced illness, injury, bereavement, stress within last 2 years. Individuals reported their education qualifications including college/university degree; A levels, AS levels, or equivalent; O levels, GCSEs, or equivalent; CSEs or equivalent; NVQ, HND, HNC, or equivalent; or other professional qualifications, or none of the above. Individuals who reported their education qualifications “none of the above” were categorized into low education level. Cumulative unfavorable SDOH were calculated based on the number of unfavorable SDOH domains (range, 0-9), and were dichotomized as 0, 1-2, 3 or more.

### Assessment of dementia

In this study, outcomes of interest were the incidence of dementia, including all-cause, and Alzheimer disease and vascular dementia. In UK Biobank, incident cases of dementia were ascertained from the primary care data and hospital inpatient records under the International Classification of Diseases, 10th revision (ICD-10) after baseline assessment. We defined outcomes according to algorithmically derived dementia outcomes (fields 42018, 42020, 42022, and 42024). Individuals were followed up from baseline (2006-2010) until the date of first diagnosis of dementia, date of death, date of loss to follow-up, the last date of hospital admission, whichever came first.

### Covariates

In this study, covariates included demographic characteristics, lifestyle factors, and clinical information. Demographic characteristics included age at baseline, gender, and race/ethnicity. Race/ethnicity status was categorized into white and other groups. In the UK Biobank, the White group included British, Irish and any other white background; the other group included Chinese, Mixed, Indian, Pakistani, Bangladeshi, Caribbean, African, and other/unknown backgrounds^28^. Lifestyle factors included current smoking (yes/no) and alcohol consumption (ever/never) status. Clinical information was obtained from self-reports, including the presence of hypertension, diabetes mellitus, heart disease, and cancer. The apolipoprotein E (APOE) genotypes were determined by a combination variant of rs429358 and rs7412, which were directly genotyped on the UK Biobank arrays^29^. Based on the number of APOE ε4 alleles, individuals were divided into APOE ε4 carriers with possessing 1 or 2 ε4 alleles; while those without were classified as APOE ε4 noncarriers^30–32^.

### Statistical Analysis

First, continuous and categorical data are presented as the means and standard deviations (SDs) or frequencies (%), respectively. The ANOVA and χ2 test were used to test the differences among categories for continuous and categorical variables, respectively. Second, we utilized Cox proportional hazard models to estimate the hazard ratios (HRs) and 95% confidence intervals (CIs) of CKM syndrome stages and SDOH associated with incident dementia. Third, we further conducted Cox proportional hazard models to estimate joint association of SDOH and CKM syndrome stages with incident dementia. All analyses were used three models. Model 1: adjustment model; Model 2: Adjusted for age, gender, body mass index, race, smoking status, and alcohol consumption. Model 3: Adjusted for Model 2 and additional APOE ε4 status. Last, we conducted Kaplan-Meier survival analysis to plot the associations of CKM syndrome stages and SDOH with incident dementia, respectively. All results were considered significant at a P value < 0.05 (2-tailed). All analyses were conducted using R statistical software (version 4.2.1; www.r-project.org).

## Results

### Characteristics of study individuals

**Table 1** shows the baseline demographic characteristics, lifestyle factors, clinical biomarkers and chronic diseases in this study. In detail, a total of 382427 individuals aged 37-73 years in UK Biobank were analyzed. Among them, 46.4% individuals were males, and 89.0% were white. The mean age of individuals was 56.50 (8.08) years. At baseline, 62147 (16.25%), 67425 (17.63%), 158637 (41.48%), 71780 (18.77%), and 22438 (5.87%) individuals were defined as CKM stage 0-4, respectively. For SODH, 37938 (10.01%) individuals were not owning a home, 30097 (7.92%) individuals were unemployment, 74487 (22.51%) had lower income, 69994 (18.34%) lived alone, 115840 (30.35%) reported less social/leisure activity, 54381 (14.68%) had fewer social supports, 31874 (8.37%) had less social connection, 170276 (44.72%) had emotional distress, 63994 (16.85%) had lower educational levels. The mean cumulative unfavorable SDOH were 1.70 (1.41). During a median follow-up period of 13.75 (IQR: 13.00-14.42) years, A total of 7174 all-cause dementia were documented, including 3201 Alzheimer’s dementia and 1131 Vascular dementia.

**Table 1.** Characteristics of study population according to CKM syndrome stage.

| **Characteristics** | **All** | **CKM Stage 0** | **CKM Stage 1** | **CKM Stage 2** | **CKM Stage 3** | **CKM Stage 4** | **P-value** |
| --- | --- | --- | --- | --- | --- | --- | --- |
| Sample, N (%) | 382427 | 62147 (16.25) | 67425 (17.63) | 158637 (41.48) | 71780 (18.77) | 22438 (5.87) |  |
| Age, Mean (SD) | 56.54 (8.08) | 53.34 (7.81) | 53.73 (7.73) | 54.86 (7.38) | 64.12 (4.31) | 61.52 (6.28) | < 0.001 |
| Gender, N (%) |  |  |  |  |  |  |  |
| Male | 205130 (53.64) | 46198 (74.34) | 46190 (68.51) | 85554 (53.93) | 19984 (27.84) | 7204 (32.11) | < 0.001 |
| Female | 177297 (46.36) | 15949 (25.66) | 21235 (31.49) | 73083 (46.07) | 51796 (72.16) | 15234 (67.89) |  |
| Race, N (%) |  |  |  |  |  |  | < 0.001 |
| White | 352593 (92.20) | 59597 (95.90) | 60081 (89.11) | 145180 (91.52) | 67134 (93.53) | 20601 (91.81) |  |
| Others | 29834 (7.80) | 2550 (4.10) | 7344 (10.89) | 13457 (8.48) | 4646 (6.47) | 1837 (8.19) |  |
| Alcohol consumption, N (%) |  |  |  |  |  |  | < 0.001 |
| Never | 29787 (7.79) | 3776 (6.08) | 4589 (6.81) | 12625 (7.96) | 6021 (8.39) | 2776 (12.37) |  |
| Ever | 352640 (92.21) | 58371 (93.92) | 62836 (93.19) | 146012 (92.04) | 65759 (91.61) | 19662 (87.63) |  |
| Current Smoking, N (%) |  |  |  |  |  |  | < 0.001 |
| No | 342744 (89.62) | 56794 (91.39) | 62372 (92.51) | 144319 (90.97) | 59715 (83.19) | 19544 (87.10) |  |
| Yes | 39683 (10.38) | 5353 (8.61) | 5053 (7.49) | 14318 (9.03) | 12065 (16.81) | 2894 (12.90) |  |
| APOE ε4 status, N (%) |  |  |  |  |  |  | < 0.001 |
| Non-carriers | 276489 (72.30) | 45011 (72.43) | 49530 (73.46) | 113530 (71.57) | 52440 (73.06) | 15978 (71.21) |  |
| Carriers | 105938 (27.70) | 17136 (27.57) | 17895 (26.54) | 45107 (28.43) | 19340 (26.94) | 6460 (28.79) |  |
| Chronic diseases, N (%) |  |  |  |  |  |  |  |
| Hypertension | 91338 (23.88) | 0 (0.00) | 0 (0.00) | 55351 (34.89) | 35536 (49.51) | 451 (2.01) | < 0.001 |
| Stroke | 6218 (1.63) | 0 (0.00) | 0 (0.00) | 0 (0.00) | 0 (0.00) | 6218 (27.71) | < 0.001 |
| Heart disease | 17239 (4.51) | 0 (0.00) | 0 (0.00) | 0 (0.00) | 0 (0.00) | 17239 (76.83) | < 0.001 |
| Diabetes | 19450 (5.09) | 0 (0.00) | 0 (0.00) | 5106 (3.22) | 10683 (14.88) | 3661 (16.32) | < 0.001 |
| Chronic kidney disease | 3743 (0.98) | 0 (0.00) | 0 (0.00) | 872 (0.55) | 2012 (2.80) | 859 (3.83) | < 0.001 |
| Overweight/Obesity | 260262 (68.06) | 0 (0.00) | 61472 (91.17) | 122876 (77.46) | 57393 (79.96) | 18521 (82.54) | < 0.001 |
| Abdominal obesity | 126231 (33.01) | 0 (0.00) | 19509 (28.93) | 66135 (41.69) | 30027 (41.83) | 10560 (47.06) | < 0.001 |
| **SDOH, N (%)** |  |  |  |  |  |  |  |
| Not owning a home | 37938 (10.01) | 4447 (7.22) | 5874 (8.79) | 16736 (10.65) | 7001 (9.83) | 3880 (17.46) | < 0.001 |
| Unemployment | 30097 (7.92) | 5001 (8.10) | 5027 (7.51) | 13756 (8.73) | 3289 (4.61) | 3024 (13.57) | < 0.001 |
| Low income | 74487 (22.51) | 8161 (14.96) | 9762 (16.56) | 28128 (20.35) | 20526 (34.02) | 7910 (42.09) | < 0.001 |
| Living alone | 69994 (18.34) | 10799 (17.41) | 11110 (16.51) | 28596 (18.06) | 14491 (20.23) | 4998 (22.34) | < 0.001 |
| Less social/leisure activity | 115840 (30.35) | 17511 (28.22) | 19198 (28.53) | 50082 (31.63) | 21660 (30.24) | 7389 (33.02) | < 0.001 |
| Fewer social support | 54381 (14.68) | 6543 (10.79) | 8015 (12.24) | 22844 (14.85) | 12399 (17.98) | 4580 (21.22) | < 0.001 |
| Less social connection | 31874 (8.37) | 4873 (7.86) | 5229 (7.78) | 13704 (8.67) | 6010 (8.41) | 2058 (9.22) | < 0.001 |
| Emotional distress | 170276 (44.72) | 26098 (42.13) | 30713 (45.74) | 73624 (46.62) | 27634 (38.68) | 12207 (54.72) | < 0.001 |
| Lower education | 63994 (16.85) | 5251 (8.48) | 8050 (12.00) | 23044 (14.62) | 20456 (28.82) | 7193 (32.44) | < 0.001 |
| Unfavorable SDOH, M (SD) | 1.70 (1.41) | 1.43 (1.25) | 1.53 (1.28) | 1.71 (1.40) | 1.86 (1.48) | 2.37 (1.69) | < 0.001 |
| Unfavorable SDOH, N (%) |  |  |  |  |  |  | < 0.001 |
| 0 | 75635 (19.78) | 14965 (24.08) | 14476 (21.47) | 30485 (19.22) | 13212 (18.41) | 2497 (11.13) |  |
| 1-2 | 215807 (56.43) | 36826 (59.26) | 40064 (59.42) | 90359 (56.96) | 37831 (52.70) | 10727 (47.81) |  |
| ≥ 3 | 90985 (23.79) | 10356 (16.66) | 12885 (19.11) | 37793 (23.82) | 20737 (28.89) | 9214 (41.06) |  |

SDOH, social determinants of health; M, mean; SD, standard deviations; APOE, apolipoprotein E.

### Association of CKM syndrome stages with incident dementia

The associations of CKM syndrome stages with dementia risk were presented in **Table 2**. After adjustment for demographic characteristics, and lifestyles, compared with individuals in CKM stage 0, the HRs for all-cause dementia were 1.00 (95% CI: 0.85-1.18), 1.14 (95% CI: 1.02-1.27), 1.18 (95% CI: 1.05-1.32), and 2.11 (95% CI: 1.82-2.44) for individuals in stage 0, 1, 2, 3 and 4, respectively. The significant greater risk for incident all-cause dementia was still observed among individuals in CKM stages 3 (HR 1.16, 95% CI: 1.02-1.31) and 4 (HR 2.03, 95% CI: 1.75-2.35) when additional adjusting for APOE ε4 status. Similarly, the higher risk of vascular dementia was also found among individuals in CKM stages 3 (HR 1.73, 95% CI: 1.27-2.35) and 4 (HR 4.66, 95% CI: 3.35-6.48) after fully adjusted for confounders, including demographic characteristics, lifestyle factors, chronic diseases, and APOE ε4 status. In addition, individuals in CKM stages 4 (HR 1.44, 95% CI: 1.15-1.81), rather than stage 1 (HR 0.83, 95% CI: 0.65-1.04), stage 2 (HR 1.10, 95% CI: 0.94-1.28) or stage 3 (HR 1.03, 95% CI: 0.87-1.22), had significantly increased risk of Alzheimer’s dementia in the fully adjusted models. Kaplan-Meier survival plots demonstrated that individuals in CKM stage 3 and 4 had significantly increased risk of incident dementia than those in stage 0 (**Figure 1**).

**Figure 1.**
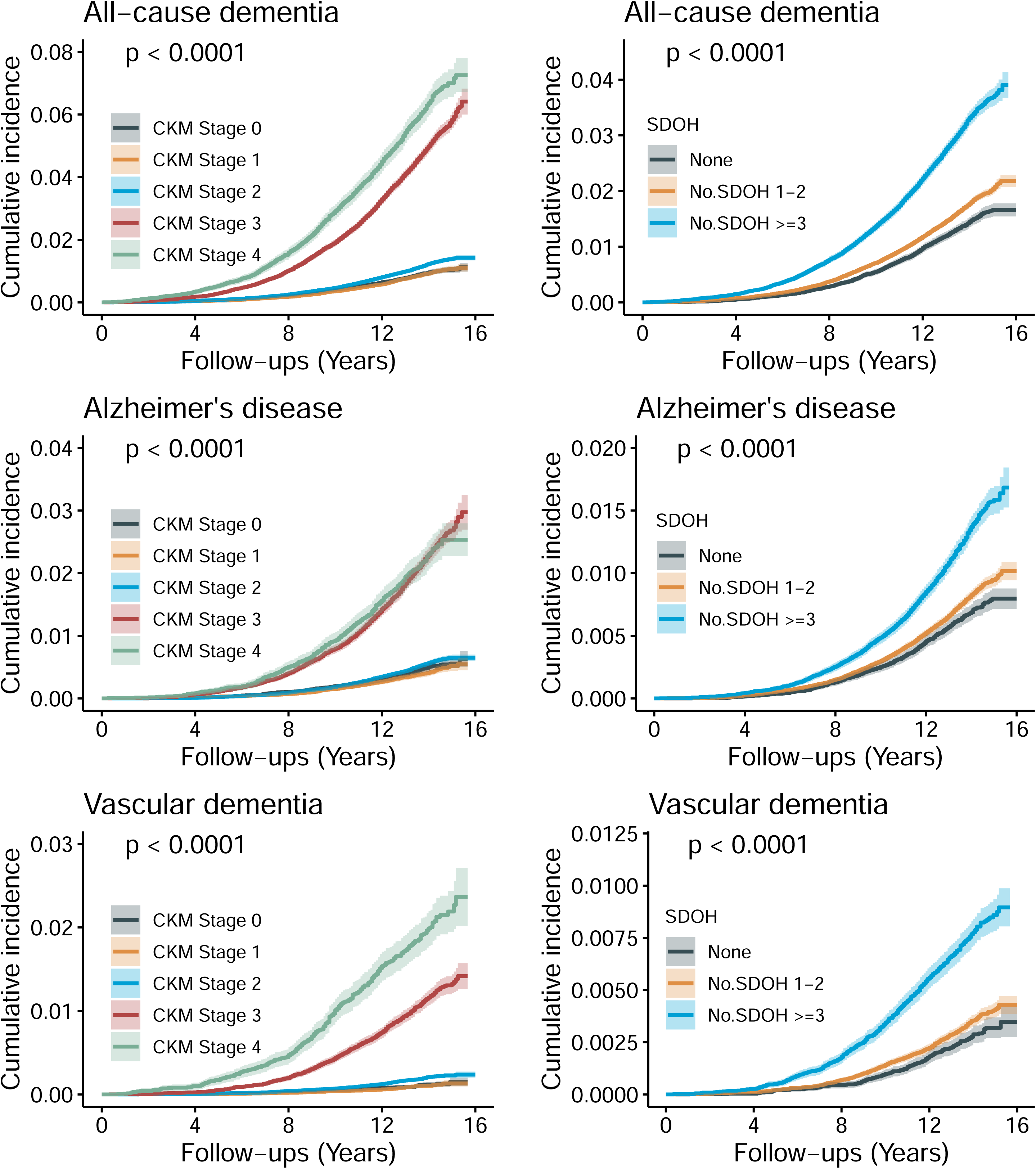
Cumulative incidence of dementia stratified by CKM syndrome stages and SDOH.

**Table 2.**
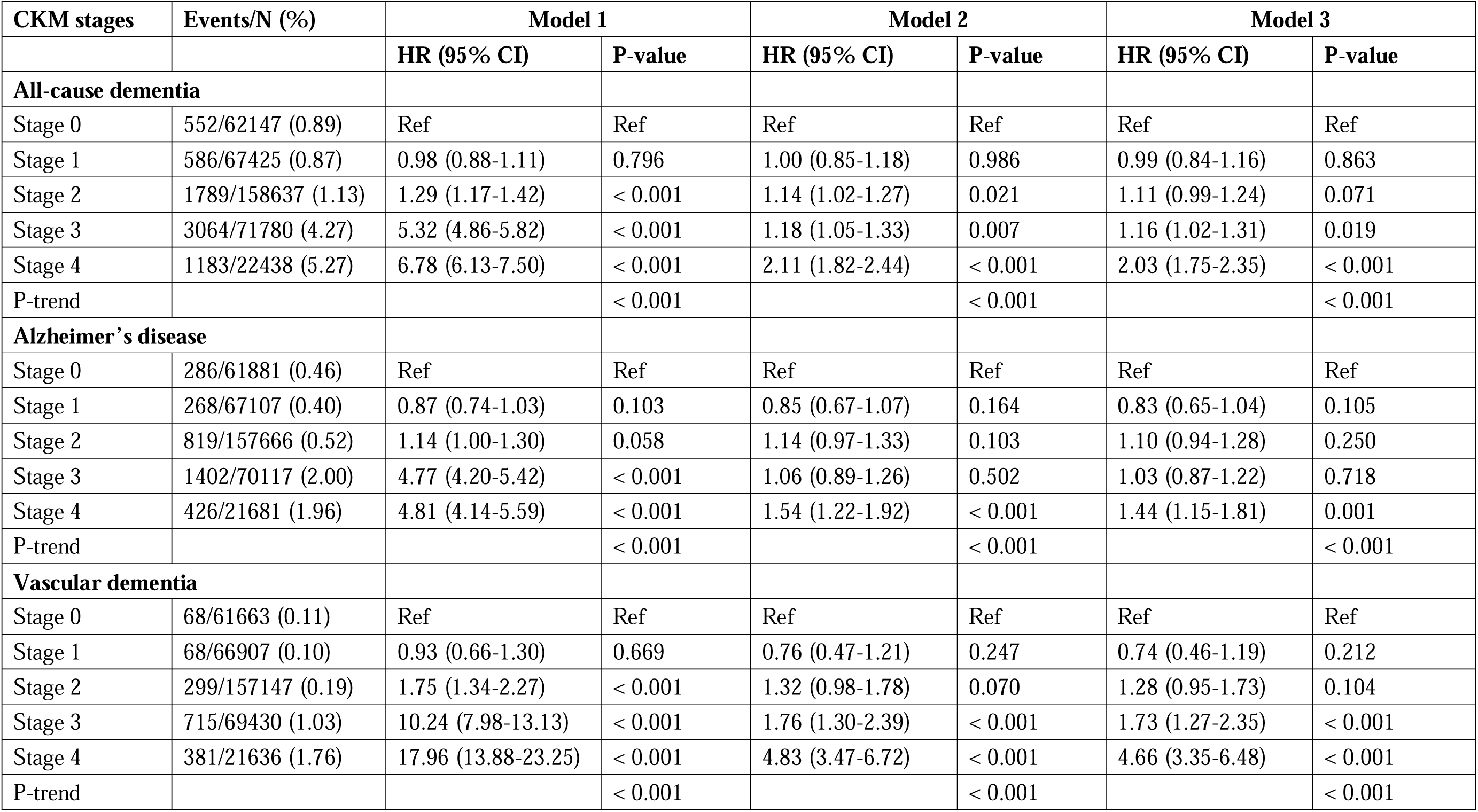
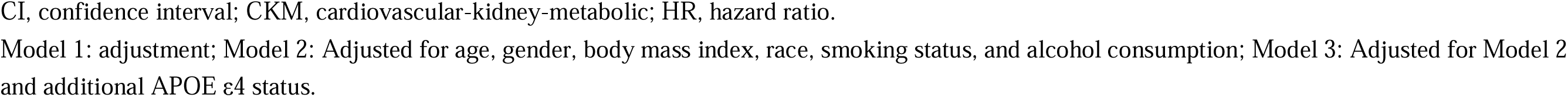
Association of stages of CKM syndrome with risk of dementia.

| CKM stages | Events/N (%) | Model 1 |  | Model 2 |  | Model 3 |  |
| --- | --- | --- | --- | --- | --- | --- | --- |
|  |  | HR (95% CI) | P-value | HR (95% CI) | P-value | HR (95% CI) | P-value |
| **All-cause dementia** |  |  |  |  |  |  |  |
| Stage 0 | 552/62147 (0.89) | Ref | Ref | Ref | Ref | Ref | Ref |
| Stage 1 | 586/67425 (0.87) | 0.98 (0.88-1.11) | 0.796 | 1.00 (0.85-1.18) | 0.986 | 0.99 (0.84-1.16) | 0.863 |
| Stage 2 | 1789/158637 (1.13) | 1.29 (1.17-1.42) | < 0.001 | 1.14 (1.02-1.27) | 0.021 | 1.11 (0.99-1.24) | 0.071 |
| Stage 3 | 3064/71780 (4.27) | 5.32 (4.86-5.82) | < 0.001 | 1.18 (1.05-1.33) | 0.007 | 1.16 (1.02-1.31) | 0.019 |
| Stage 4 | 1183/22438 (5.27) | 6.78 (6.13-7.50) | < 0.001 | 2.11 (1.82-2.44) | < 0.001 | 2.03 (1.75-2.35) | < 0.001 |
| P-trend |  |  | < 0.001 |  | < 0.001 |  | < 0.001 |
| **Alzheimer's disease** |  |  |  |  |  |  |  |
| Stage 0 | 286/61881 (0.46) | Ref | Ref | Ref | Ref | Ref | Ref |
| Stage 1 | 268/67107 (0.40) | 0.87 (0.74-1.03) | 0.103 | 0.85 (0.67-1.07) | 0.164 | 0.83 (0.65-1.04) | 0.105 |
| Stage 2 | 819/157666 (0.52) | 1.14 (1.00-1.30) | 0.058 | 1.14 (0.97-1.33) | 0.103 | 1.10 (0.94-1.28) | 0.250 |
| Stage 3 | 1402/70117 (2.00) | 4.77 (4.20-5.42) | < 0.001 | 1.06 (0.89-1.26) | 0.502 | 1.03 (0.87-1.22) | 0.718 |
| Stage 4 | 426/21681 (1.96) | 4.81 (4.14-5.59) | < 0.001 | 1.54 (1.22-1.92) | < 0.001 | 1.44 (1.15-1.81) | 0.001 |
| P-trend |  |  | < 0.001 |  | < 0.001 |  | < 0.001 |
| **Vascular dementia** |  |  |  |  |  |  |  |
| Stage 0 | 68/61663 (0.11) | Ref | Ref | Ref | Ref | Ref | Ref |
| Stage 1 | 68/66907 (0.10) | 0.93 (0.66-1.30) | 0.669 | 0.76 (0.47-1.21) | 0.247 | 0.74 (0.46-1.19) | 0.212 |
| Stage 2 | 299/157147 (0.19) | 1.75 (1.34-2.27) | < 0.001 | 1.32 (0.98-1.78) | 0.070 | 1.28 (0.95-1.73) | 0.104 |
| Stage 3 | 715/69430 (1.03) | 10.24 (7.98-13.13) | < 0.001 | 1.76 (1.30-2.39) | < 0.001 | 1.73 (1.27-2.35) | < 0.001 |
| Stage 4 | 381/21636 (1.76) | 17.96 (13.88-23.25) | < 0.001 | 4.83 (3.47-6.72) | < 0.001 | 4.66 (3.35-6.48) | < 0.001 |
| P-trend |  |  | < 0.001 |  | < 0.001 |  | < 0.001 |

CI, confidence interval; CKM, cardiovascular-kidney-metabolic; HR, hazard ratio.
Model 1: adjustment; Model 2: Adjusted for age, gender, body mass index, race, smoking status, and alcohol consumption; Model 3: Adjusted for Model 2 and additional APOE ε4 status.

### Association of SODH with incident dementia

In this study, we examined the association between SODH and risk of incident dementia (**Table 3**). After adjusting for demographic characteristics, lifestyles, and APOE ε4 status, we found that increasing numbers of unfavorable SDOH were dose-dependently associated with an increased risk of all-cause dementia (HR 1.20, 95% CI: 1.18-1.22). Compared with individuals without unfavorable SDOH, those with 1-2 (HR 1.24, 95% CI: 1.16-1.33) and ≥3 (HR 1.87, 95% CI: 1.73-2.01) unfavorable SDOH had higher risk of incident all-cause dementia. Kaplan-Meier survival plots also demonstrated similar results (**Figure 1**). Unfavorable SDOH was both associated with increased risk of all-caused dementia in the fully adjusted models (**Supplemental Table 2**), including not owning a home (HR 1.84, 95% CI: 1.72-1.98), unemployment (HR 2.03, 95% CI: 1.85-2.23), lower income (HR 1.59, 95% CI: 1.51-1.68), living alone (HR 1.29, 95% CI: 1.23-1.37), less social/leisure activity (HR 1.18, 95% CI: 1.12-1.24), fewer social support (HR 1.19, 95% CI: 1.12-1.26), less social connection (HR 1.27, 95% CI: 1.17-1.37), emotional distress (HR 1.20, 95% CI: 1.14-1.25), and lower educational levels (HR 1.40, 95% CI: 1.33-1.47). Additionally, similar findings were observed in the association of unfavorable SODH with Alzheimer’s disease and vascular dementia, with HRs ranging from 1.11 to 2.44.

**Table 3.** Association of SDOH with risk of dementia.

| **SDOH** | **Events/N (%)** | **Model 1** |  | **Model 2** |  | **Model 2** |  |
| --- | --- | --- | --- | --- | --- | --- | --- |
|  |  | **HR (95% CI)** | **P-value** | **HR (95% CI)** | **P-value** | **HR (95% CI)** | **P-value** |
| **All-cause dementia** |  |  |  |  |  |  |  |
| Per additional factor | 7174/382427 (1.88) | 1.25 (1.23-1.27) | < 0.001 | 1.20 (1.18-1.22) | < 0.001 | 1.20 (1.18-1.22) | < 0.001 |
| SDOH category |  |  |  |  |  |  |  |
| 0 | 1019/75635 (1.35) | Ref | Ref | Ref | Ref | Ref | Ref |
| 1-2 | 3524/215807 (1.63) | 1.23 (1.14-1.32) | < 0.001 | 1.24 (1.16-1.33) | < 0.001 | 1.24 (1.16-1.33) | < 0.001 |
| ≥ 3 | 2631/90985 (2.89) | 2.27 (2.12-2.44) | < 0.001 | 1.86 (1.72-2.00) | < 0.001 | 1.87 (1.73-2.01) | < 0.001 |
| **Alzheimer's disease** |  |  |  |  |  |  |  |
| Per additional factor | 3201/378452 (0.86) | 1.19 (1.17-1.22) | < 0.001 | 1.15 (1.12-1.18) | < 0.0001 | 1.16 (1.13-1.18) | < 0.001 |
| SDOH category |  |  |  |  |  |  |  |
| 0 | 487/75103 (0.65) | Ref | Ref | Ref | Ref | Ref | Ref |
| 1-2 | 1630/213912 (0.76) | 1.19 (1.07-1.32) | < 0.001 | 1.21 (1.09-1.34) | < 0.001 | 1.21 (1.09-1.34) | < 0.001 |
| ≥ 3 | 1084/89437 (1.21) | 1.98 (1.78-2.20) | < 0.001 | 1.64 (1.47-1.83) | < 0.001 | 1.65 (1.48-1.85) | < 0.001 |
| **Vascular dementia** |  |  |  |  |  |  |  |
| Per additional factor | 1531/376783 (0.41) | 1.32 (1.28-1.36) | < 0.001 | 1.25 (1.21-1.29) | < 0.0001 | 1.25 (1.21-1.29) | < 0.001 |
| SDOH category |  |  |  |  |  |  |  |
| 0 | 198/74814 (0.26) | Ref | Ref | Ref | Ref | Ref | Ref |
| 1-2 | 703/212986 (0.66) | 1.26 (1.08-1.48) | 0.004 | 1.26 (1.08-1.48) | 0.004 | 1.26 (1.08-1.48) | 0.004 |
| ≥ 3 | 630/88983 (0.71) | 2.82 (2.40-3.31) | < 0.001 | 2.20 (1.86-2.59) | < 0.001 | 2.21 (1.88-2.61) | < 0.001 |

SDOH: social determinants of health; CI, confidence interval; CKM, cardiovascular-kidney-metabolic; HR, hazard ratio.
Model 1: adjustment; Model 2: Adjusted for age, gender, body mass index, race, smoking status, and alcohol consumption; Model 3: Adjusted for Model 2 and additional APOE ε4 status.

### Joint association of CKM syndrome stages and SODH with incident dementia

The joint association of CKM syndrome stages and SODH with risk of incident dementia were presented in **Table 4**. Compared with the reference group (individuals in CKM stage 0 and no unfavorable SDOH), individuals with ≥ 3 unfavorable SDOH had higher risks of incident all-cause dementia in CKM syndrome stage 0 (HR 1.61, 95% CI: 1.25-2.07), stage 1 (HR 1.38, 95% CI: 1.00-1.91), stage 2 (HR 1.76, 95% CI: 1.41-2.21), stage 3 (HR 1.90, 95% CI: 1.50-2.42), and stage 4 (HR 3.63, 95% CI: 2.80-4.71), respectively. Meanwhile, individuals with 1-2 unfavorable SDOH showed higher risks of incident all-cause dementia in CKM syndrome stage 2 (HR 1.31, 95% CI: 1.06-1.62), stage 3 (HR 1.33, 95% CI: 1.06-1.68), and stage 4 (HR 2.14, 95% CI: 1.64-2.77), respectively. Similar results were observed in the association of CKM syndrome stages and SODH with incident Alzheimer’s and vascular dementia. Individuals with ≥ 3 unfavorable SDOH and CKM syndrome stage 4 also had the highest risk of incident Alzheimer’s disease (HR 2.22, 95% CI: 1.47-3.37) and vascular dementia (HR 7.42, 95% CI: 4.20-13.09), respectively.

**Table 4.** Joint associations of SDOH and CKM syndrome with incident dementia.

|  | Events/N | Model 1 |  | Model 2 |  | Model 3 |  |
| --- | --- | --- | --- | --- | --- | --- | --- |
|  |  | HR (95% CI) | P-value | HR (95% CI) | P-value | HR (95% CI) | P-value |
| **All-cause dementia** |  |  |  |  |  |  |  |
| CKM Stage 0 |  |  |  |  |  |  |  |
| SDOH = 0 | 113/14965 (0.76) | Ref | Ref | Ref | Ref | Ref | Ref |
| SDOH 1-2 | 294/36826 (0.80) | 1.06 (0.86-1.32) | 0.573 | 1.11 (0.89-1.38) | 0.359 | 1.11 (0.90-1.38) | 0.333 |
| SDOH $\geq 3$ | 145/10356 (1.40) | 1.91 (1.49-2.44) | < 0.001 | 1.58 (1.23-2.03) | < 0.001 | 1.61 (1.25-2.07) | < 0.001 |
| CKM Stage 1 |  |  |  |  |  |  |  |
| SDOH=0 | 101/14476 (0.70) | 0.93 (0.71-1.21) | 0.588 | 1.23 (0.85-1.78) | 0.274 | 1.23 (0.85-1.78) | 0.279 |
| SDOH 1-2 | 324/40064 (0.81) | 1.08 (0.88-1.34) | 0.459 | 1.13 (0.87-1.48) | 0.367 | 1.12 (0.86-1.47) | 0.393 |
| SDOH $\geq 3$ | 161/12885 (1.25) | 1.72 (1.35-2.18) | < 0.001 | 1.39 (1.00-1.92) | 0.048 | 1.38 (1.00-1.91) | 0.050 |
| CKM Stage 2 |  |  |  |  |  |  |  |
| SDOH = 0 | 251/30485 (0.82) | 1.10 (0.88-1.37) | 0.399 | 1.06 (0.82-1.37) | 0.668 | 1.03 (0.79-1.34) | 0.830 |
| SDOH 1-2 | 921/90359 (1.02) | 1.37 (1.13-1.67) | 0.001 | 1.34 (1.09-1.66) | 0.006 | 1.31 (1.06-1.62) | 0.011 |
| SDOH $\geq 3$ | 617/37793 (1.63) | 2.26 (1.85-2.76) | < 0.001 | 1.79 (1.43-2.25) | < 0.001 | 1.76 (1.41-2.21) | < 0.001 |
| CKM Stage 3 |  |  |  |  |  |  |  |
| SDOH = 0 | 455/13212 (3.44) | 4.92 (4.00-6.04) | < 0.001 | 1.04 (0.78-1.38) | 0.813 | 1.03 (0.77-1.38) | 0.827 |
| SDOH 1-2 | 1449/37831 (3.83) | 5.61 (4.63-6.79) | < 0.001 | 1.37 (1.09-1.72) | 0.007 | 1.33 (1.06-1.68) | 0.013 |
| SDOH $\geq 3$ | 1160/20737 (5.59) | 8.66 (7.13-10.50) | < 0.001 | 1.93 (1.52-2.45) | < 0.001 | 1.90 (1.50-2.42) | < 0.001 |
| CKM Stage 4 |  |  |  |  |  |  |  |
| SDOH = 0 | 99/2497 (3.96) | 5.78 (4.41-7.57) | < 0.001 | 1.34 (0.88-2.03) | 0.167 | 1.26 (0.83-1.91) | 0.277 |
| SDOH 1-2 | 536/10727 (5.00) | 7.43 (6.06-9.10) | < 0.001 | 2.23 (1.72-2.89) | < 0.001 | 2.14 (1.64-2.77) | < 0.001 |
| SDOH $\geq 3$ | 548/9214 (5.95) | 9.44 (7.71-11.56) | < 0.001 | 3.60 (2.77-4.68) | < 0.001 | 3.56 (2.73-4.62) | < 0.001 |
| **Alzheimer's disease** |  |  |  |  |  |  |  |
| CKM Stage 0 |  |  |  |  |  |  |  |
| SDOH=0 | 55/14907 (0.37) | Ref | Ref | Ref | Ref | Ref | Ref |
| SDOH 1-2 | 163/36695 (0.44) | 1.21 (0.89-1.65) | 0.216 | 1.26 (0.93-1.71) | 0.141 | 1.27 (0.94-1.73) | 0.123 |
| SDOH ≥ 3 | 68/10279 (0.66) | 1.85 (1.30-2.64) | < 0.001 | 1.59 (1.11-2.29) | 0.012 | 1.64 (1.14-2.36) | 0.007 |
| CKM Stage 1 |  |  |  |  |  |  |  |
| SDOH=0 | 49/14424 (0.34) | 0.93 (0.63-1.36) | 0.695 | 1.20 (0.71-2.03) | 0.505 | 1.20 (0.70-2.03) | 0.508 |
| SDOH 1-2 | 154/39894 (0.39) | 1.06 (0.78-1.44) | 0.709 | 0.96 (0.65-1.40) | 0.815 | 0.94 (0.64-1.39) | 0.763 |
| SDOH ≥ 3 | 65/12789 (0.51) | 1.43 (1.00-2.05) | 0.050 | 1.12 (0.68-1.83) | 0.651 | 1.12 (0.68-1.83) | 0.656 |
| CKM Stage 2 |  |  |  |  |  |  |  |
| SDOH=0 | 131/30365 (0.43) | 1.18 (0.86-1.62) | 0.301 | 1.19 (0.82-1.71) | 0.363 | 1.15 (0.80-1.65) | 0.463 |
| SDOH 1-2 | 421/89858 (0.47) | 1.29 (0.98-1.71) | 0.074 | 1.45 (1.07-1.96) | 0.016 | 1.40 (1.03-1.89) | 0.030 |
| SDOH ≥ 3 | 267/37443 (0.71) | 2.02 (1.51-2.70) | < 0.001 | 1.79 (1.29-2.48) | < 0.001 | 1.76 (1.27-2.44) | < 0.001 |
| CKM Stage 3 |  |  |  |  |  |  |  |
| SDOH=0 | 214/12971 (1.65) | 4.80 (3.57-6.45) | < 0.001 | 1.02 (0.67-1.55) | 0.926 | 1.02 (0.67-1.55) | 0.913 |
| SDOH 1-2 | 680/37062 (1.83) | 5.48 (4.16-7.21) | < 0.001 | 1.45 (1.05-2.01) | 0.024 | 1.41 (1.02-1.95) | 0.039 |
| SDOH ≥ 3 | 508/20084 (2.53) | 7.97 (6.03-10.53) | < 0.001 | 1.49 (1.05-2.10) | 0.024 | 1.47 (1.04-2.07) | 0.028 |
| CKM Stage 4 |  |  |  |  |  |  |  |
| SDOH=0 | 38/2436 (1.56) | 4.63 (3.06-7.00) | < 0.001 | 1.63 (0.89-2.97) | 0.113 | 1.53 (0.84-2.81) | 0.164 |
| SDOH 1-2 | 212/10403 (2.04) | 6.15 (4.57-8.27) | < 0.001 | 1.77 (1.20-2.62) | 0.004 | 1.66 (1.13-2.46) | 0.010 |
| SDOH ≥ 3 | 176/8842 (1.99) | 6.41 (4.73-8.68) | < 0.001 | 2.27 (1.50-3.44) | < 0.001 | 2.22 (1.47-3.37) | < 0.001 |
| **Vascular dementia** |  |  |  |  |  |  |  |
| CKM Stage 0 |  |  |  |  |  |  |  |
| SDOH=0 | 18/14870 (0.12) | Ref | Ref | Ref | Ref | Ref | Ref |
| SDOH 1-2 | 27/36559 (0.07) | 0.61 (0.34-1.12) | 0.109 | 0.63 (0.34-1.14) | 0.126 | 0.64 (0.35-1.16) | 0.140 |
| SDOH ≥ 3 | 23/10234 (0.22) | 1.91 (1.03-3.53) | 0.040 | 1.44 (0.76-2.73) | 0.265 | 1.46 (0.77-2.78) | 0.244 |
| CKM Stage 1 |  |  |  |  |  |  |  |
| SDOH=0 | 8/14383 (0.06) | 0.46 (0.20-1.06) | 0.068 | 0.64 (0.21-1.98) | 0.437 | 0.63 (0.20-1.96) | 0.426 |
| SDOH 1-2 | 35/39775 (0.09) | 0.74 (0.42-1.30) | 0.290 | 0.69 (0.33-1.45) | 0.329 | 0.69 (0.33-1.44) | 0.320 |
| SDOH ≥ 3 | 25/12749 (0.20) | 1.67 (0.91-3.06) | 0.097 | 1.25 (0.55-2.83) | 0.600 | 1.22 (0.54-2.78) | 0.629 |
| CKM Stage 2 |  |  |  |  |  |  |  |
| SDOH=0 | 46/30280 (0.15) | 1.27 (0.73-2.18) | 0.396 | 0.99 (0.52-1.89) | 0.984 | 0.97 (0.51-1.85) | 0.923 |
| SDOH 1-2 | 151/89589 (0.17) | 1.42 (0.87-2.31) | 0.163 | 1.18 (0.70-2.00) | 0.538 | 1.15 (0.68-1.95) | 0.596 |
| SDOH ≥ 3 | 102/37278 (0.27) | 2.35 (1.43-3.88) | < 0.001 | 1.82 (1.04-3.19) | 0.037 | 1.79 (1.02-3.14) | 0.042 |
| CKM Stage 3 |  |  |  |  |  |  |  |
| SDOH=0 | 92/12849 (0.72) | 6.35 (3.83-10.53) | < 0.001 | 0.90 (0.46-1.79) | 0.769 | 0.90 (0.45-1.78) | 0.756 |
| SDOH 1-2 | 336/36718 (0.92) | 8.28 (5.15-13.31) | < 0.001 | 1.59 (0.93-2.74) | 0.093 | 1.56 (0.91-2.69) | 0.106 |
| SDOH ≥ 3 | 287/19863 (1.44) | 13.65 (8.48-21.98) | < 0.001 | 3.09 (1.76-5.42) | < 0.001 | 3.06 (1.75-5.37) | < 0.0001 |
| CKM Stage 4 |  |  |  |  |  |  |  |
| SDOH=0 | 34/2432 (1.40) | 12.52 (7.07-22.17) | < 0.001 | 3.23 (1.41-7.38) | 0.005 | 2.97 (1.29-6.82) | 0.010 |
| SDOH 1-2 | 154/10345 (1.49) | 13.59 (8.34-22.15) | < 0.001 | 4.00 (2.24-7.17) | < 0.001 | 3.87 (2.16-6.93) | < 0.001 |
| SDOH ≥ 3 | 193/8859 (2.18) | 21.12 (13.03-34.24) | < 0.001 | 7.48 (4.24-13.19) | < 0.001 | 7.42 (4.20-13.09) | < 0.001 |

SDOH: social determinants of health; CI, confidence interval; CKM, cardiovascular-kidney-metabolic; HR, hazard ratio.
Model 1: adjustment; Model 2: Adjusted for age, gender, body mass index, race, smoking status, and alcohol consumption; Model 3: Adjusted for Model 2 and additional APOE ε4 status.

## Discussion

In this large-scale, population-based, and prospective cohort study from the UK Biobank, we revealed dose-response associations of unfavorable SDOH and advanced CKM syndrome with increased risk of incident dementia, independently of demographic characteristics, lifestyle factors, and APOE ε4 status. Significantly higher dementia risks were also found in individuals with any unfavorable SDOH, such as not owning a home, unemployment, and living alone. In addition, we also observed a joint association of CKM syndrome stages and SODH with incident dementia. Notably, individuals with ≥3 unfavorable SDOH and in CKM syndrome stage 4 had the highest risk of incident all-cause, Alzheimer’s disease and vascular dementia.

At present, CKM syndrome was recently introduced by the AHA, and was reported to be association with increased risks of chronic diseases and mortality in diverse populations^12–15^. However, fewer study was conducted to examined the association between CKM syndrome stage and dementia risk^15^. Jiang et.al included 2641 participants age 70-79 (mean age 74 ± 2.8) years in the Health, Aging, and Body Composition study, with a 15-year follow-up^15^. They found that participants with CKM stages 3-4 had a 50% increase in dementia risk (HR 1.50, 95 % CI: 1.20-1.86) than those in stage 0-2 in the fully adjusted model, which is consistence with our findings. Obviously, they just adjusted for demographic characteristics and lifestyles on the association of CKM syndrome stage with dementia risk, but not considered the effect of genetic predisposition. They also lacked data on the etiology of dementia, such as Alzheimer’s disease and vascular dementia^15^. In addition, the limited sample size and elderly-focused population may introduce bias, and potentially skew the observed association between CKM syndrome and dementia risk.

In our study, using large-scale and population-based data from UK Biobank, we took further steps of examining the association of CKM syndrome with dementia. We found that CKM syndrome stage 3 and 4 was associated with increased risk of incident all-cause dementia, as well as Alzheimer’s disease and vascular dementia, independently of demographic characteristics, lifestyles, and APOE ε4 status. Our findings provided robust evidence of the momentous effect of CKM syndrome on the development of dementia. In addition, previous studies reported statistically significant disparities in the prevalence of CKM stages by SDOH [13-16], and significant association between unfavorable SDOH and dementia risk^2–5^. For example, Zhu et.al conducted a study to examine the prevalence of CKM syndrome across SODH using data from the National Health and Nutrition Examination Survey (1999-2018) ^20^. They found that, compared with their favorable counterparts, higher prevalence of CKM syndrome stage 4 was observed in individuals with unemployment (14.0% [95% CI: 12.9%-15.2] vs 6.8% [95% CI: 6.4%-7.2%]), and lower education (11.5% [95% CI: 10.6%-12.5%] vs 7.4% [95% CI: 6.9%-7.8%]). Meanwhile, Zhang et.al found significant association of unemployment (HR 1.96, 95% CI: 1.72-2.24), and lower education (HR 1.17, 95% CI: 1.10-1.23) with increased risk of dementia using data from UK Biobank^2^. These findings highlighted the potential role of SODH on the relationship between CKM syndrome stage and dementia risk. Therefore, we further examined the joint association of CKM syndrome and SDOH with dementia risk in this study, and found an additive effect of CKM syndrome and cumulative unfavorable SDOH on the risk of developing dementia. In summary, to our knowledge, this study is the first to report the associations of CKM syndrome stage in different numbers of SDOH for dementia risk, and underscore the urgent need to address these inequities to help prevent dementia.

### Strengths and Limitations

Our study has several strengths. We conducted this study in a large-scale, population-based, and prospective cohort study with longer follow-up periods. This study allowed us to examine the associations between CKM syndrome stage, SODH, and the incidence of dementia with enhanced statistical power and precision. The robustness of our findings, bolstered by the comprehensive nature of our data, suggests that our results may be more generalizable. We considered effects of genetic predisposition (APOE ε4 status) on these associations, and also released consistent findings. Additionally, this study represented the first investigation into the associations between CKM syndrome stage and dementia risk across different numbers of SDOH factors. Our findings could provide valuable information to identify vulnerable populations at an increased risk of dementia. Limitations in this study should also be noted. First, as this was an observational study, the associations of CKM syndrome stage and SODH with incident dementia were merely correlational rather than causal. Second, CKM syndrome stage and SODH were assessed only at one time point, and it might change in the follow-up. Therefore, we did not capture its trajectories over time, and might lead to potential misclassification in this study. Finally, although we controlled for many potential confounders, we cannot rule out all potential residual confounders, especially unmeasured variables.

## Conclusion

In this large-scale, population-based, and prospective cohort study from the UK Biobank, we evaluated different CKM syndrome stage and SODH, and revealed significant associations of unfavorable SDOH and advanced CKM syndrome with increased risks of incident dementia, independently of demographic characteristics, lifestyle factors, and APOE ε4 status. Novelty, we also observed a joint association of CKM syndrome stages and unfavorable SODH with incident dementia. Our findings highlighted the significance of both maintaining optimal CKM health and addressing unfavorable SODH in the context of cognitive aging. Further studies involving diverse populations and assessing a broader range of SODH are warranted to confirm our findings and elucidate the underlying mechanisms.

## Data Availability

Data from UK Biobank are available on application at www.ukbiobank.ac.uk/register-apply.

## Acknowledgements

The data used in this research were obtained from the UK Biobank. We would like to thank the workers, researchers, and individuals involved in the UK Biobank. This research has been conducted using the UK Biobank Resource under Application Number 103791.

## Data sharing statement

Data from UK Biobank are available on application at www.ukbiobank.ac.uk/register-apply.

## Author contributions

Dr. Meng Hao and Hui Zhang had full access to all the data in the study and takes responsibility for the integrity of the data and the accuracy of the data analysis.

Concept and design: Hui Zhang, Meng Hao, Zixin Hu, Shuai Jiang, and Xiangwei Li. Acquisition, analysis, or interpretation of data: Xiangnan Li, Shuming Wang, Meng Hao. Drafting of the manuscript: Hui Zhang, Meng Hao.

Critical revision of the manuscript for important intellectual content: All authors. Statistical analysis: Meng Hao, Xiangnan Li, Shuai Jiang.

Administrative, technical, or material support: Xiangnan Li, Xiangwei Li, Xiaofeng Wang, and Yi Li.

Supervision: Meng Hao, Shuai Jiang, Xiangwei Li.

---

**Supplementary Table 1. Assessment of SDOH in UK Biobank**.

**Supplemental Table 2. Association of individual SDOH with risk of dementia**.

