## Supplemental Tables for "Cardiovascular-Kidney-Metabolic Syndrome, Social Determinants of Health, and Dementia Risk: a prospective cohort study"

**Supplementary Table 1. Assessment of SDOH in UK Biobank.**

| **Social determinant of health** | **Questionnaires** | **Definition of unfavorable SDOH** | **Fields** |
| --- | --- | --- | --- |
| Total household income | "What is the average total income before tax received by your HOUSEHOLD?" | 0: Average total household income before tax is more than £18000  1: Less than £18000 | 738 |
| Employment status | "Which of the following describes your current situation? | 0: In paid employment or self-employed; Retired; Full or part-time student  1: Unemployed; Looking after home and/family; Unable to work because of sickness or disability; Doing unpaid or voluntary work; None of above | 6142 |
| Home ownership | "Do you own or rent the accommodation that you live in?" | 0: Own outright; Own with a mortgage  1: Rent | 680 |
| Living status | "Including yourself, how many people are living together in your household?” | 0: Number in household is more than 1  1: Number in household is 1 | 709 |
| Social support | "How often are you able to confide in someone close to you?" | 0: Almost daily; 2-4 times a week; About once a week; About once a month; Once every few month  1: Never or almost never | 2110 |
| Social activity | "Which of the following do you attend once a week or more often? " | 0: Sports club or gym; Pub or social club; Religious group; Adults’ education class; Other group activity  1: None of the above | 6160 |
| Social isolation | "How often do you visit friends or family or have them visit you?" | 0: Almost daily; 2-4 times a week; About once a week; About once a month;  1: Once every few months; Never or almost never; No friends/family outside household | 1031 |
| Emotional distress | "In the last 2 years have you experienced any of the following? " | 0: Serious illness, injury or assault to yourself; Serious illness, injury or assault of a close relative; Death of a close relative; Death of a spouse or partner; Marital separation/divorce; Financial difficulties  1: None of the above | 6145 |
| Educational levels | "Which of the following qualifications do you have?” | 0: College/university degree, and others. Individuals who reported their education qualifications including college/university degree; A levels, AS levels, or equivalent; O levels, GCSEs, or equivalent; CSEs or equivalent; NVQ, HND, HNC, or equivalent; or other professional qualifications,  1: None of the above | 6138 |

**Supplemental Table 2. Association of individual SDOH with risk of dementia.**

| **SDOH** | **Events/N (%)** | **Model 1** |  | **Model 2** |  | **Model 2** |  |
| --- | --- | --- | --- | --- | --- | --- | --- |
|  |  | **HR (95% CI)** | **P-value** | **HR (95% CI)** | **P-value** | **HR (95% CI)** | **P-value** |
| **All-cause dementia** | |  |  |  |  |  |  |
| Not owning a home | 998/37938 (2.63) | 1.55 (1.45-1.65) | 3.27E-37 | 1.84 (1.72-1.97) | 2.58E-67 | 1.84 (1.72-1.98) | 1.91E-67 |
| Unemployment | 509/30097 (1.69) | 0.92 (0.84-1.01) | 0.077283787 | 2.05 (1.87-2.25) | 4.16E-51 | 2.03 (1.85-2.23) | 6.06E-50 |
| Lower income | 2516/74487 (3.38) | 2.93 (2.78-3.09) | 0 | 1.59 (1.50-1.68) | 1.09E-61 | 1.59 (1.51-1.68) | 3.47E-62 |
| Living alone | 1806/69994 (2.58) | 1.56 (1.48-1.65) | 6.29E-60 | 1.29 (1.23-1.37) | 9.07E-21 | 1.29 (1.23-1.37) | 1.02E-20 |
| Less social/leisure activity | 2261/115840 (1.95) | 1.07 (1.02-1.13) | 0.005233012 | 1.18 (1.12-1.24) | 9.57E-11 | 1.18 (1.12-1.24) | 1.63E-10 |
| Fewer social support | 1299/54381 (2.39) | 1.37 (1.29-1.46) | 5.12E-25 | 1.18 (1.11-1.25) | 1.24E-07 | 1.19 (1.12-1.26) | 3.03E-08 |
| Less social connection | 634/31874 (1.99) | 1.09 (1.01-1.18) | 0.034795315 | 1.26 (1.16-1.37) | 4.02E-08 | 1.27 (1.17-1.37) | 1.95E-08 |
| Emotional distress | 3064/170276 (1.80) | 0.95 (0.90-0.99) | 0.021828606 | 1.19 (1.14-1.25) | 2.95E-13 | 1.20 (1.14-1.25) | 1.37E-13 |
| Lower education | 2416/63994 (3.78) | 2.68 (2.55-2.81) | 0 | 1.39 (1.32-1.47) | 7.47E-38 | 1.40 (1.33-1.47) | 2.42E-38 |
| **Alzheimer’s dementia** | |  |  |  |  |  |  |
| Not owning a home | 403/37343 (1.08) | 1.39 (1.25-1.54) | 9.74E-10 | 1.77 (1.59-1.97) | 2.33E-25 | 1.78 (1.60-1.98) | 8.00E-26 |
| Unemployment | 170/29758 (0.57) | 0.68 (0.58-0.79) | 8.66E-07 | 1.72 (1.47-2.02) | 2.39E-11 | 1.70 (1.45-2.00) | 5.94E-11 |
| Lower income | 1089/73060 (1.49) | 2.96 (2.73-3.21) | 1.28E-156 | 1.54 (1.41-1.67) | 4.74E-24 | 1.54 (1.42-1.68) | 1.47E-24 |
| Living alone | 752/68939 (1.09) | 1.43 (1.32-1.55) | 9.94E-18 | 1.15 (1.06-1.25) | 0.00069934 | 1.15 (1.06-1.25) | 0.000716181 |
| Less social/leisure activity | 986/114564 (0.86) | 1.04 (0.97-1.12) | 0.299873662 | 1.18 (1.10-1.27) | 1.53E-05 | 1.18 (1.09-1.27) | 2.15E-05 |
| Fewer social support | 528/53609 (0.98) | 1.24 (1.13-1.36) | 5.96E-06 | 1.09 (1.00-1.20) | 0.0609524 | 1.11 (1.01-1.22) | 0.03540897 |
| Less social connection | 248/31488 (0.79) | 0.95 (0.83-1.08) | 0.4092667 | 1.16 (1.01-1.32) | 0.029945025 | 1.17 (1.02-1.33) | 0.020892506 |
| Emotional distress | 1241/168453 (0.74) | 0.80 (0.75-0.86) | 1.54E-09 | 1.04 (0.97-1.12) | 0.3048971 | 1.04 (0.97-1.12) | 0.243144885 |
| Lower education | 1130/62707 (1.80) | 2.90 (2.70-3.12) | 1.05E-180 | 1.47 (1.37-1.59) | 2.86E-24 | 1.48 (1.37-1.59) | 8.34E-25 |
| **Vascular dementia** | |  |  |  |  |  |  |
| Not owning a home | 246/37186 (0.66) | 1.83 (1.60-2.10) | 3.31E-18 | 2.07 (1.80-2.38) | 7.12E-24 | 2.07 (1.80-2.39) | 5.50E-24 |
| Unemployment | 117/29705 (0.39) | 1.00 (0.83-1.20) | 0.977935939 | 2.45 (2.01-2.98) | 7.19E-19 | 2.42 (1.99-2.95) | 1.79E-18 |
| Lower income | 606/72577 (0.83) | 3.55 (3.18-3.97) | 7.74E-110 | 1.82 (1.62-2.04) | 7.43E-24 | 1.82 (1.62-2.05) | 3.77E-24 |
| Living alone | 387/68575 (0.56) | 1.57 (1.40-1.76) | 1.40E-14 | 1.32 (1.17-1.48) | 4.08E-06 | 1.32 (1.17-1.48) | 4.00E-06 |
| Less social/leisure activity | 510/114088 (0.45) | 1.17 (1.05-1.30) | 0.004559194 | 1.27 (1.14-1.41) | 1.47E-05 | 1.27 (1.14-1.41) | 1.69E-05 |
| Fewer social support | 298/53379 (0.56) | 1.51 (1.33-1.71) | 2.20E-10 | 1.23 (1.08-1.39) | 0.0018505 | 1.24 (1.09-1.40) | 0.001165181 |
| Less social connection | 136/31376 (0.43) | 1.10 (0.92-1.31) | 0.291135523 | 1.23 (1.03-1.47) | 0.021576 | 1.23 (1.03-1.47) | 0.020005274 |
| Emotional distress | 685/167897 (0.41) | 1.03 (0.93-1.14) | 0.618655639 | 1.30 (1.18-1.44) | 2.97E-07 | 1.31 (1.18-1.45) | 2.51E-07 |
| Lower education | 550/62127 (0.89) | 3.01 (2.71-3.34) | 4.80E-94 | 1.43 (1.29-1.60) | 5.68E-11 | 1.44 (1.29-1.60) | 3.19E-11 |

**Model 1:** adjustment; **Model 2:** Adjusted for age, gender, body mass index, race, smoking status, and alcohol consumption; **Model 3:** Adjusted for Model 2 and additional APOE ε4 status.
